# Antigenic and virological properties of an H3N2 variant that will likely dominate the 2021-2022 Northern Hemisphere influenza season

**DOI:** 10.1101/2021.12.15.21267857

**Authors:** Marcus J. Bolton, Jordan T. Ort, Ryan McBride, Nicholas J. Swanson, Jo Wilson, Moses Awofolaju, Allison R. Greenplate, Elizabeth M. Drapeau, Andrew Pekosz, James C. Paulson, Scott E. Hensley

## Abstract

Influenza viruses have circulated at very low levels during the COVID-19 pandemic, and population immunity against these viruses is low. Influenza virus cases have been increasing in the Northern Hemisphere involving an H3N2 strain (3C.2a1b.2a2) with a hemagglutinin (HA) that has several substitutions relative to the 2021-2022 H3N2 vaccine strain. Here, we show that one of these substitutions eliminates a key glycosylation site on HA and alters sialic acid binding. Using glycan array profiling, we show that the 3C.2a1b.2a2 H3 maintains binding to an extended bi-antennary sialoside and replicates to high titers in human airway cells. We found that antibodies elicited by the 2021-2022 Northern Hemisphere influenza vaccine poorly neutralize the new H3N2 strain. Together, these data indicate that 3C.2a1b.2a2 H3N2 viruses efficiently replicate in human cells and could potentially cause an antigenic mismatch if they continue to circulate at high levels during the 2021-2022 influenza season.

## Main

Population immunity against influenza viruses is likely low since these viruses have not circulated widely during the COVID-19 pandemic^1^. Social distancing, mask wearing, and decreases in international travel have likely contributed to reduced global circulation of influenza viruses^2^. Once COVID-19-related restrictions are eased or lifted, it is possible that influenza viruses will circulate widely due to lack of infection-induced population immunity over the past two years. In recent weeks, a unique H3N2 clade, 3C.2a1b.2a2 (herein 2a2), has circulated at elevated levels in the United States and other parts of the world (**Figure 1A**). This clade emerged early in the COVID-19 pandemic and almost completely displaced other H3N2 clades in Europe, Oceania, South Asia, West Asia, and North America in 2021. Viruses within this clade have several substitutions in key antigenic sites on hemagglutinin (HA) relative to the 2021-2022 Northern Hemisphere H3N2 vaccine strain, a 3C.2a1b.2a1 (herein 2a1) virus.

**Figure 1.**
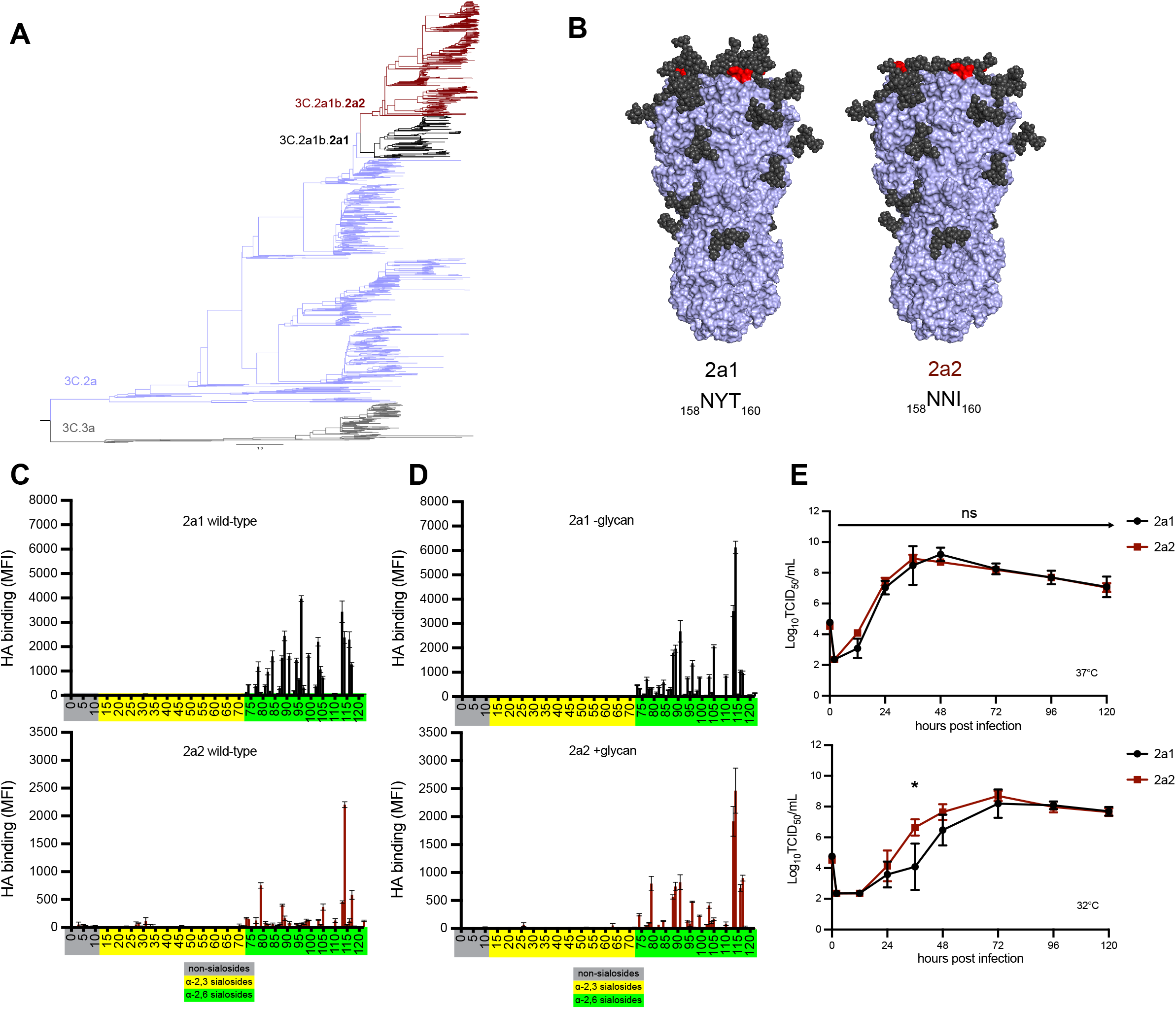
H3N2 variant lineage 2a2 replicates efficiently in human cells despite changes in HA receptor specificity. **a** Nextstrain phylogenetic analysis of the HA gene of contemporary (2019-2021) H3N2 viruses. **b** Amino acid differences at residues 158-160 are denoted (red) and glycan loss is modeled on the crystal structure of an H3 HA trimer (PDB 4O5I) for 2a1 and 2a2 HAs **c**,**d** Binding of wild-type (**c**) and mutant (**d**) recombinant HAs to a sialoside microarray. Printed glycans include those with α2,3 sialic acid linkages (yellow), α2,6 sialic acid linkages (green), or no sialic acid linkage (grey). Bars represent the mean fluorescent intensity with standard error bars. **e** Infectious virus production after an MOI=0.01 infection of primary differentiated human nasal epithelial cell (hNEC) cultures incubated at 37°C (top) or 32°C (bottom). A repeated measures MANOVA followed by a Bonferonni post-test was completed (^*^p≤0.05).

Vaccine effectiveness against 2a2 was found to be low during a recent outbreak on a college campus in the United States during November 2021^3^. It is therefore important to understand the virological and antigenic properties of 2a2 H3N2 viruses.

Viruses within the 2a2 H3N2 clade possess a T160I HA substitution that is expected to eliminate a glycosylation motif at amino acids 158-160 in antigenic site B (**Figure 1B**). Recombinant HA proteins engineered to possess I160 (2a2 and 2a1-glycan) migrated with a lower molecular weight compared to HAs with T160 (2a1 and 2a2 +glycan) (**Extended Data 1**), consistent with a loss of a glycosylation site with the T160I substitution. Influenza virus entry into human cells is mediated by HA binding to α2,6-linked sialoglycans^4^, and previous studies have demonstrated that the 158-160 HA glycosylation site impacts HA sialic acid binding specificity^5^. To determine if the 2a2 H3N2 HA has altered receptor specificity, we measured HA binding to a panel of α2,3- and α2,6-linked sialoside glycans (**Figure 1C, Supplementary Table 1**). As expected, HAs from the predecessor 2a1 H3N2 virus and the new 2a2 H3N2 virus did not bind strongly to α2,3-linked sialosides or those lacking sialic acid altogether (non-sialosides).

Consistent with previous studies^6^, the predecessor 2a1 H3N2 HA bound to many α2,6-linked sialoside glycans, with preference to those with extended poly-LacNAc chains and/or branched structures. Strikingly, the new 2a2 H3N2 HA bound poorly to most α2,6-linked sialoside glycans, but maintained strong binding to an extended bi-antennary sialoside. We also measured the binding of 2a1 and 2a2 mutant HAs which we generated via site-directed mutagenesis to either remove (in the case of 2a1) or add (in the case of 2a2) a glycosylation motif at positions 158-160 (**Figure 1D**). Interestingly, mutant 2a1 HA that lacked the 158-160 glycan (2a1-glycan) had a similar binding profile to the wild-type 2a2 HA, with a strong preference for an extended bi-antennary sialoside (#114, **Supplementary Table 1**). Conversely, the 2a2 HA engineered to possess the 158-160 glycan (2a2 +glycan) bound with broader specificity than the wild-type 2a2 HA to multiple α2,6-linked sialoside glycans (**Figure 1E**). These data suggest a role for the 158-160 glycan in shaping the binding specificity of the new 2a2 H3N2 HAs for sialoside receptors.

The types and prevalence of different sialo-linked receptors in the human respiratory tract is incompletely understood, and therefore it is difficult to interpret how changes in sialoside glycan specificity may affect viral fitness. We measured viral growth kinetics over 120 hours in primary human nasal epithelial cells (hNECs) (**Figure 1F**) and found that both 2a1 and 2a2 H3N2 viruses replicated well in hNEC cultures at 37°C, with no significant differences in infectious virus peak titers or kinetics of virus production between the two viruses (**Figure 1F, top**). When we measured growth kinetics of the two viruses in hNEC cultures incubated at 32°C, which is a temperature more consistent with that of the upper respiratory tract, we again found that both viruses replicated well, but at this lower temperature the new 2a2 H3N2 virus replicated with slightly faster kinetics than 2a1 H3N2, though both viruses reached the same peak titer at the same time (**Figure 1F, bottom**). Additionally, both viruses replicated to similar levels in MDCK-S and hCK transformed cell lines engineered to support human-adapted influenza virus replication (**Extended Data 1**). These data indicate that 2a2 H3N2 viruses, despite a decrease in receptor specificity breadth, replicate at least as efficiently as 2a1 H3N2 viruses in both primary and transformed cell culture systems.

We previously demonstrated that the addition of a 158-160 H3 glycan resulted in a major antigenic change during the 2014-2015 season^7,8^, which likely contributed to low vaccine effectiveness that season^9^. Most influenza vaccines are based on egg-adapted influenza virus antigens, and we previously reported that mutations at HA residues 158-160 can occur as a result of egg-adaptation^8^. To determine if the new 2a2 H3N2 virus is antigenically distinct from the 2021-2022 Northern Hemisphere H3N2 component, we completed neutralization assays using serum collected from 40 individuals before and after receiving the egg-adapted Flulaval Quadrivalent vaccine. We tested antibody neutralization of wild-type and egg-adapted 2a1 H3N2 strains, as well as the new 2a2 H3N2 strain. Importantly, the wild-type 2a1 HA encodes amino acids Asn, Tyr, Thr at positions 158-160 which leads to a glycosylation site, whereas the egg-adapted 2a1 HA encodes amino acids Asn, Tyr, Lys at positions 158-160 which abrogates this glycosylation site (**Figure 2A**). We found that most individuals possessed antibodies that neutralized the egg-adapted 2a1 strain but not the wild-type version of this strain or the new 2a2 strain (**Figure 2B-C**). Neutralizing antibodies against the egg-adapted vaccine strain were higher relative to the other strains tested before vaccination (**Figure 2B**) and were boosted ∼2 fold after vaccination (**Figure 2C**). Neutralizing antibodies against wild-type 2a1 and the new 2a2 strain were very low before vaccination and remained undetectable in many individuals (35% to wild-type 2a1; 55% to 2a2) following vaccination (**Figure 2C**). On average, neutralizing antibody titers were reduced ∼10 fold comparing the new 2a2 H3N2 strain relative to the egg-adapted 2a1 vaccine strain (**Figure 2D**), which is likely an underestimation since samples with an undetectable neutralization titer were arbitrarily assigned a neutralization titer of 10 (the limit of detection in our assays).

**Figure 2.**
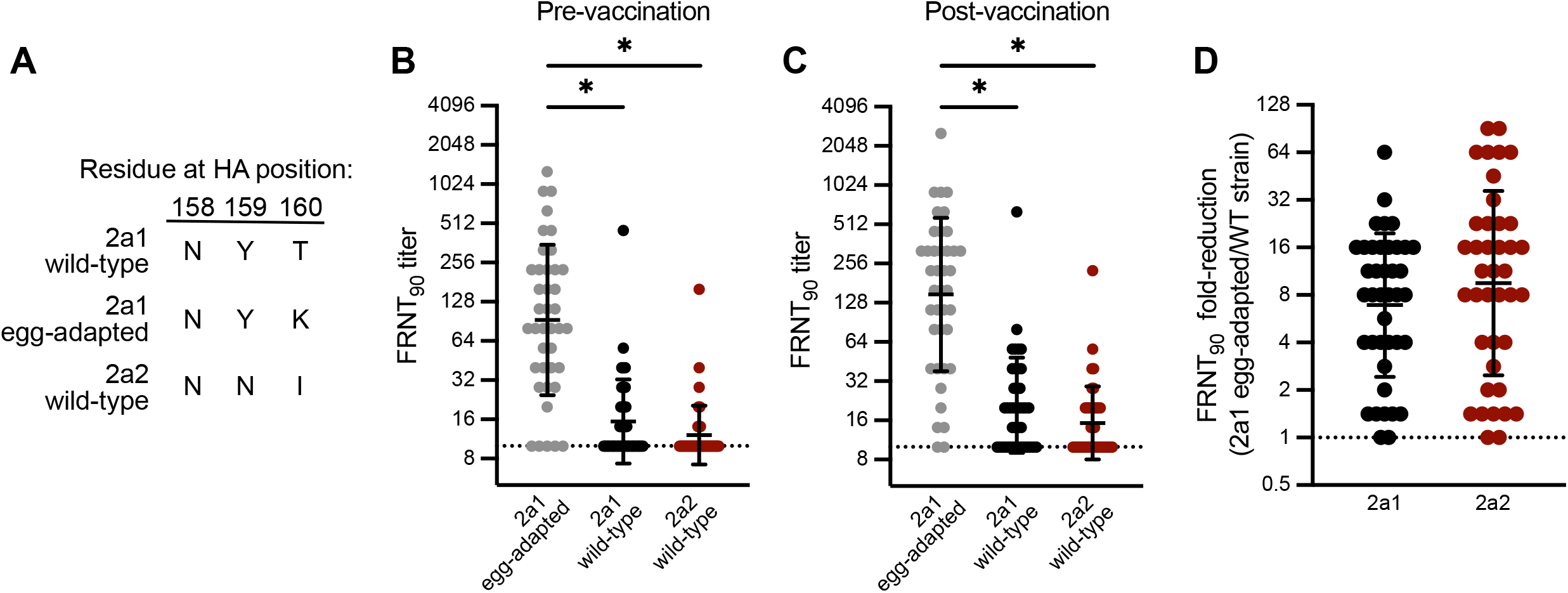
Potential H3N2 antigenic mismatch for the 2021-2022 Northern Hemisphere influenza season. **a** Amino acid residues encoded by egg-adapted and wild-type circulating strains at HA positions 158, 159, and 160, an N-linked glycosylation site. **b**,**c** Virus neutralization titers to the vaccine virus and circulating viruses prior to vaccination (**b**) and 27-28 days following vaccination (**c**) in a group of vaccinees in Oct-Nov 2021 (n=40). Neutralization titers are plotted as the geometric mean titer (GMT) of two independent experiments. Lines represent the GMT and standard deviation among vaccinees. A Kruskal-Wallis one-way ANOVA with Dunn’s multiple comparisons test was completed on log_2_ transformed titers (^*^p≤0.05). **d** Fold-reduction in neutralizing antibodies to circulating strains when compared to the vaccine virus in a group of vaccinees (n=40)

Further studies will be needed to evaluate antibody responses elicited by other vaccine types that employ wild-type 2a1 H3N2 strains, such as cell-based or recombinant protein vaccines^10^. While there is still a potential antigenic mismatch between wild-type 2a1 HA and the new 2a2 HA, it is possible that glycan shielding from the 158-160 glycan on wild-type vaccine antigens may promote antibody responses directed against epitopes other than antigenic site B of HAs. It is worth noting that most vaccinees in our study were relatively young and healthy and additional studies will need to be completed with larger cohorts to better evaluate the 2021-2022 Northern Hemisphere vaccine in individuals with different birth years.

Our studies suggest that the new 2a2 H3N2 replicates efficiently in human airway cells and can partially circumvent antibodies elicited by egg-adapted 2021-2022 Northern Hemisphere influenza vaccines. We were hopeful that the egg-adapted H3N2 vaccine strain would be closely matched to the new 2a2 H3N2 strain since both of these strains lack a glycan at residues 158-160 of HA. However, our data indicate that these H3N2 strains are antigenically mismatched, likely due to amino acid differences at HA residues 159 and 160. The new 2a2 H3N2 strain lost the 158-160 antigenic site B glycan through a T160I substitution, while the egg-adapted 2021-2022 H3N2 strain lost it through a T160K substitution. Previous studies have demonstrated that amino acid differences at HA residues 158-160 can impact antigenicity^11^, and it is likely that the amino acid differences at residue 160 contribute to this vaccine mismatch.

While cases of 2a2 H3N2 infections are quickly rising in the United States and other parts of the world, it is possible that other clades of H3N2 will become predominant in the future. It is also possible that H1N1 or influenza B viruses might dominate later in the 2021-2022 season.

Studies have clearly shown that seasonal influenza vaccines consistently prevent hospitalizations and deaths even in years where there are large antigenic mismatches^12^. Influenza vaccinations will be crucial for reducing hospitalizations as SARS-CoV-2 and 2a2 H3N2 viruses co-circulate in the coming months.

## Methods

### Human serum samples

Serum samples from 40 adults (ages 21-71) were collected at the time of and 27-28 days after vaccination with Flulaval Quadrivalent influenza virus vaccine (GlaxoSmithKline) between October-December 2021. This study was approved by the Institutional Review Board of the University of Pennsylvania.

### Cell lines and primary cells

MDCK-SIAT1 (MDCK-S) cells were cultured in Minimal Essential Medium (MEM) supplemented with 10% fetal bovine serum (FBS), and 293T cells were cultured in Dulbecco’s Minimal Essential Medium (DMEM) with 10% FBS. hCK cells^13^ were cultured in MEM supplemented with 10% newborn calf serum. All transformed cell lines were cultured at 37C in humidified incubators supplemented with 5% CO_2_. Primary Human Nasal Epithelial Cells (hNECs, Promocell, Heidelberg, Germany) were plated and cultured in serum-free Airway Epithelial Growth Medium (Promocell) with SupplementPack Airway Epithelial Cell Growth Medium without antibiotics. The 24 well plate Transwell inserts were coated with 0.03 mg/mL Collagen I, Rat Tail (Gibco). Cells were plated and cultured with Airway media on the apical and basolateral surface with media changes at 24 hours after cell plating and every 48 hours afterwards. After 7-10 days, confluence was assessed by light microscopy and by determining transepithelial electrical resistance (TEER). When fully confluent (TEER> 400), both apical and basolateral media were removed and ALI Differentiation media (Stem Cell Technologies, Pneumacult ALI Basal Medium supplemented with 1X ALI Maintenance Supplement (StemCell Technologies), 048 ug/mL Hydrocortisone solution (StemCell Technologies), and 4 μg/mL Heparin sodium salt in PBS (StemCell Technologies) was replaced in the basolateral chamber only. Media was changed every 48 hours and the apical surface of the cultures was intermittently washed with PBS to remove excess mucus. Full differentiation occurred in approximately 4 weeks and cells were considered fully differentiated when there was presence of mobile cilia on the cell surface visible with light microscopy.

### Viruses

Influenza viruses were generated by an 8 plasmid reverse genetics system as previously described^14^ with internal segments from A/Puerto Rico/8/1934. The wild-type HA and NA sequences for A/Cambodia/e0826360/2020 (3C.2a1b.2a1) and the HA sequence for A/Bangladesh/1450/2020 (3C.2a1b.2a2) were synthesized as gene fragments (IDT) and cloned into the pHW2000 reverse genetics plasmid by restriction digest cloning. Viruses were launched by transient transfection of 8 plasmids encoding each of the influenza virus genome segments in co-cultures of 293T and MDCK-S cells. Virus stocks were passaged one time in MDCK-S cells and aliquots were stored at −80ºC. All virus stocks were sequence confirmed by Sanger sequencing.

### Recombinant proteins

Soluble recombinant HA trimers were produced as previously described^15^. HA mammalian expression plasmids encoded for a codon-optimized HA sequence followed by a T4 fibritin FoldOn trimerization domain, an AviTag biotinylation sequence, and a 6xHistidine tag at the C-terminus in place of the transmembrane domain and cytoplasmic tail of HA. Proteins were produced by transient transfection in 293F cells (Thermo Fisher), and purified using Ni-NTA agarose beads (Qiagen) using gravity flow chromatography columns (Bio-Rad). Site-directed mutants were generated in HA expression plasmids using the QuikChange II-XL kit according to manufacturer’s instructions (Aglient). All plasmids were sequence confirmed prior to transfection, and all proteins were checked for integrity and the absence of contaminants by SDS-PAGE.

### Glycan arrays

Glycans were synthesized and printed on microarray chips as previously described^6^. Recombinant HA protein (50 μg/mL), a mouse anti-histidine tag antibody, and an Alexa647-conjugated anti-mouse antibody were pre-complexed in a 4:2:1 ratio on ice for 15 minutes in 100 μL PBS-T. Pre-complexed HAs were incubated on the microarray surface for 90 minutes in a humidified chamber.

### Virus growth curves

The hNECs were acclimated to 32ºC or 37ºC for 48hrs before infection. The apical surface was washed three times with PBS and the basolateral media was changed at the time of infection. The hNEC cultures were inoculated at an ∼MOI of 0.01 in the apical chamber and incubated at 32ºC incubator for 2 hours. The apical surface of the hNEC culture was washed three times with PBS+. At the indicated times, 100 μL of IM without N-acetyl trypsin was added to the apical surface of the hNECs for 5 minutes at 32ºC, the IM was harvested and stored at −80ºC. Basolateral media was changed every 24hrs post infection for the duration of the experiment. Infectious virus titers in the apical supernatants were measured with TCID50 assay^16^.

For growth assays in MDCK-S and hCK cells, triplicate wells of a 6-well plate were seeded with 10^6^ cells/well. 12 hours later, cells were washed 2 times with serum free MEM (SF-MEM) and then infected with virus at a multiplicity of infection of 0.0001 and incubated at 37ºC for 1 hour. Following virus adsorption, inoculum was removed and cells were washed twice with SF-MEM. MEM supplemented with 5 mM HEPES buffer, 50 μg/mL gentamycin-sulfate, and 1 μg/mL TPCK-treated trypsin (growth media) was then added to wells, and plates were incubated at 37C + 5% CO_2_. At designated times post-infection, a 400 μL aliquot was removed from each well, clarified by centrifugation, and frozen at −80ºC. Media depleted from wells was replaced with growth media.

### Focus reduction neutralization test (FRNT)

FRNT assays were completed as previously described^17^. Prior to testing, serum samples were treated with receptor destroying enzyme (RDE) (Denka-Seiken) for two hours at 37ºC and then inactivated for 30 minutes at 56ºC. On the day of the assay, serum samples were serially diluted in serum-free MEM and mixed with a virus dilution that resulted in 300-500 focus forming units (FFU) per well in virus only control wells. 96-well tissue culture plates containing MDCK-S cells plated the previous day at 25,000 cells/well were washed three times with serum-free MEM. Virus-antibody mixtures were added to washed cells and incubated for 1 hour at 37C. Virus-antibody mixtures were removed from cells, cells were washed with serum-free MEM and media was replaced with MEM supplemented with 5 mM HEPES buffer, 50 μg/mL gentamycin-sulfate, and 1.25% Avicel. Plates were incubated for 18 hours in a 37ºC incubator supplied with 5% CO_2_. For visualizing infected cells, media was removed, and cells were fixed with 4% paraformaldehyde for 1 hour at 4ºC. Fixed cells were permeabilized with 0.5% Triton X-100 for 7 minutes, and then blocked for 1 hour with 5% milk in PBS. A mouse anti-nucleoprotein antibody (clone IC5-1B7), and then a peroxidase-conjugated goat anti-mouse antibody were each diluted in blocking buffer and incubated on plates for 1 hour after each other. Plates were washed 5 times with distilled water after blocking, primary, and secondary steps. TrueBlue TMB substrate (KPL) was added to plates and incubated in the dark for 1 hour for foci development. Substrate was flicked out of wells, and plates were thoroughly dried before visualization on an ImmunoSpot S6 plate reader. FRNT_90_ titers were defined as the highest reciprocal serum dilution that inhibited ≥90% of virus, according to virus only control wells on each plate.

## Data Availability

All data produced in the present work are contained in the manuscript.

## Acknowledgments

We thank Yoshihiro Kawaoka for providing hCK cells and Sarah Cobey for providing comments on the manuscript. We thank Penn’s Immune Health for processing human samples for this study.

---

**Extended Data 1.**
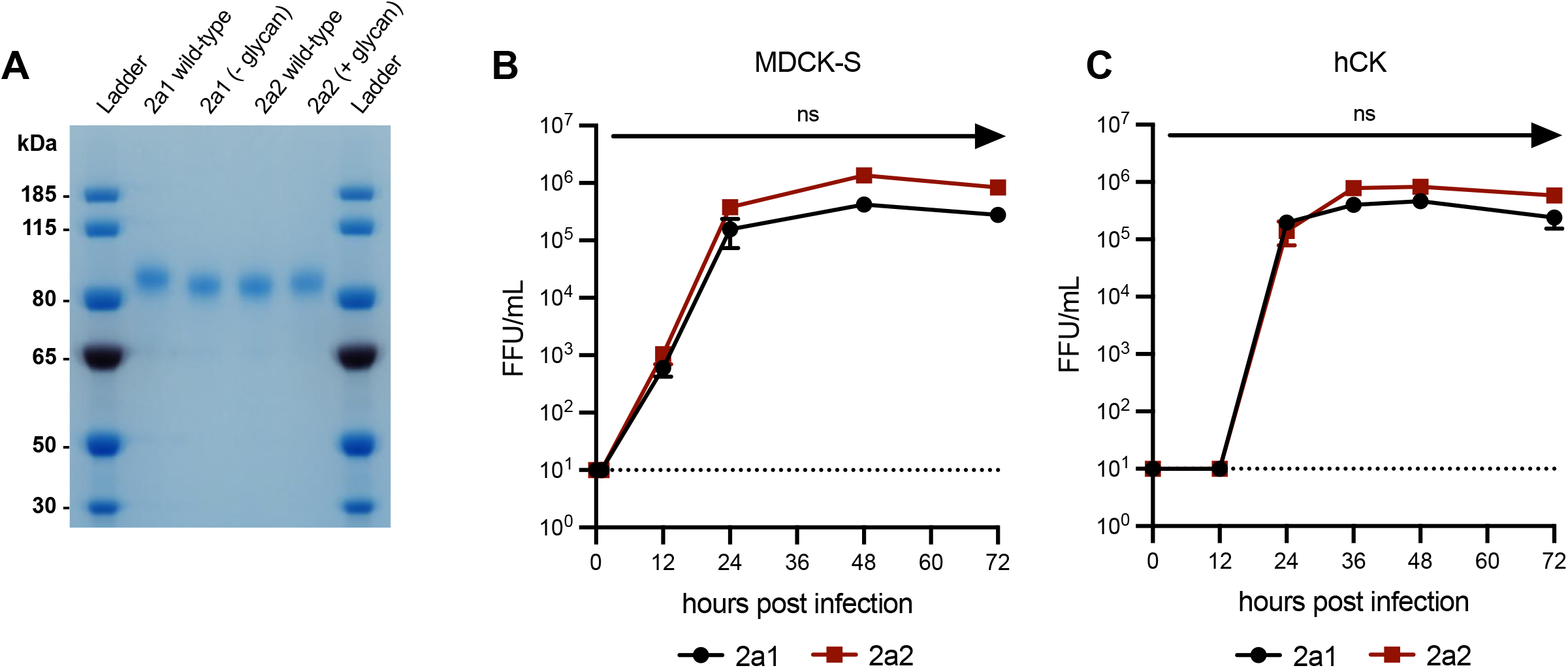
**a** SDS-PAGE analysis of wild-type and mutant recombinant HA proteins. **b**,**c** Infectious virus production following infection with virus at MOI of 0.0001 in MDCK-S (**b**) and hCK (**c**) cells incubated at 37 C. Statistical comparison of viruses was completed using a Welch’s t-test on log_10_-transformed titers at each timepoint post infection (^*^p≤0.05).

